# Eye Movements During the Iowa Gambling Task in Parkinson’s Disease: A Brief Report

**DOI:** 10.1101/2024.12.07.24318658

**Authors:** Kirby Doshier, Anthony J. Ryals, Vicki A. Nejtek, Michael F. Salvatore

## Abstract

Parkinson’s disease (PD) is characterized by motor and cognitive impairments. Subtle cognitive impairment may precede motor impairment. There is a substantial need for innovative assessments, such as those involving decision-making, to detect PD in the premotor phase. Evidence suggests executive dysfunction in PD can impede strategic decision-making relying on learning and applying feedback. The Iowa Gambling Task (IGT), when combined with eye-tracking, may be a valuable synergistic strategy for predicting impaired decision-making and therapeutic non-compliance. Participants with PD and matched healthy controls completed the Movement Disorders Society’s modified Unified Parkinson’s Disease Rating Scale (UPDRS-MDS), 6-minute Walk test (6MWT), Timed Up and Go test (TUG), Trail Making Test A and B (TMT A and B), Controlled Oral Word Association Test (COWAT), and the Barratt Impulsiveness Scale (BIS). Eye tracking was recorded during the IGT. The PD group scored significantly higher on UPDRS subscales and covered less distance during the 6MWT despite equivalent performance on the TUG. The PD group also had longer completion times on TMT A and B and more errors on TMT B. Overall IGT winning scores were marginally worse in PD. However, when analyzed as a function of performance over time, the PD group performed significantly worse by task end, thus suggesting impaired decision-making. PD participants exhibited a 72% reduction in blinks despite equivalent outcomes in other eye-movements. Combined with established motor and executive function tests, the IGT and similar tasks combined with eye-tracking may be a powerful noninvasive method to detect and monitor PD early in progression.

---

Parkinson’s disease (PD) is characterized by distinct motor dysfunctions; bradykinesia, tremors, and rigidity, that are associated with degeneration of dopaminergic neurons in the substantia nigra leading to a loss of dopamine signaling (1–3). Importantly, non-motor symptoms occur in roughly 90% of PD patients (4) with a prodromal period including depression, anxiety, and cognitive difficulties preceding motor symptoms (5). Accordingly, up to 50% of PD patients experience a form of mild cognitive impairment characterized by subtle cognitive changes not always apparent on standardized tests (6) These changes may directly affect quality of life including through therapeutic non-compliance (7, 8). Some have proposed that cognitive deficits can be present up to 10 years before a diagnosis of PD occurs (9). Because the subtleties in cognitive dysfunction in PD are under-detected and under-treated in clinical practice, it is highly relevant for clinicians to be skilled in recognizing the cognitive domains that are most at risk for early decline. A need exists for techniques combining decision-making behavior with psychophysiological correlates, such as eye tracking, to better characterize and diagnose early PD progression (10).

Individuals with PD can experience progressive changes in executive functioning contributing to goal-directed behavior and adjustments to novel situations (11,12). The prefrontal cortex (PFC) is critically involved in executive functioning, such as through selection and inhibition (13–15), conflict monitoring (16) and risk and reward processing (17). Individuals with PD have demonstrated impaired evaluation of rewarding outcomes (18), a tendency toward risk taking, and an inability to learn from negative feedback (19).

The Iowa Gambling Task (IGT) is one of the most widely used assessments for understanding decision making abilities under conditions of reward and uncertainty in individuals with PD (19, 20). Salvatore and colleagues propose that the IGT may be sensitive to detecting prodromal changes due to its close relationship with several key regions involved in the neurological dysfunction of PD patients (e.g., frontal cortex, locus coeruleus) (20). While the existing literature is mixed, PD patients often score significantly lower than healthy controls on the IGT task (e.g., 21-22), perhaps reflecting an inability to balance reward and punishment (23–24). Neurobiologically, this may be due to a significant loss of dopaminergic midbrain functions (e.g., substantia nigra and ventral tegmental area) and noradrenergic function through the locus coeruleus which in turn impairs connections to the prefrontal cortex (20,25,26). Accordingly, some have estimated a 40-77% loss of dopamine-based innervation in the ventral tegmentum and a 63% cell loss in norepinephrine-based cell loss in locus coeruleus in patients with PD (25–26).

Eye movements (fixations, saccades, blinks, and pupil dilation) are robust reflections of both perceptual and cognitive functions (27), and multiple mid-brain and frontal connections are involved in eye movements (28–30). For instance, dopaminergic activity involved in learning, memory, and goal-oriented behavior can be indirectly measured through spontaneous eyeblink rate during visual exploration (31). Specifically, dopamine is thought to modulate the frequency of these spontaneous eyeblinks involved in reward-driven behavior and cognitive flexibility (32). Eyeblinks are also related to dopaminergic activity in basal ganglia by contributing to both motor and cognitive functioning (33,34). Pupil dilation is mediated by locus coeruleus in response to autonomic arousal and cognitive functioning (35). The locus coeruleus system releases norepinephrine which has widespread effects on central and peripheral nervous system. Degeneration of the LC is considered a critical component of PD because it contributes to cognitive impairment and motor dysfunction (36). Understanding the relationship between cognitive deficits and abnormalities of eye movements offers promise as a marker of early neurodegeneration (37). Unfortunately, limited research exists linking decision-making with eye movements in prodromal PD (37–41). To our knowledge, no study to date has used the IGT as a platform for measuring eye movements during decision making in individuals with PD.

## The Present Study

We report outcomes of tests of motor functioning and cognitive control in a sample of 25 early-stage PD patients evaluated during their “ON” state and 20 age-matched and education-matched healthy controls. PD patients presented at a mean Hoehn and Yahr Stage of 1.44 indicating unilateral and unliteral/axial involvement (42). Motor functioning was assessed using the Unified Parkinson’s Disease Rating Scale (43), a 6-minute Walk Test, a Timed Up and Go (TUG) test, and a gait speed measurement. Verbal fluency was assessed using the controlled oral word association test (COWAT; 44), impulsivity was assessed using the Barratt’s Impulsiveness Scale (BIS; 45), and cognitive flexibility/working memory was assessed using the Trail Making A and B tasks (46). Participants completed a computerized of the Iowa Gambling Task (47) while simultaneously measuring eye movements. We first hypothesized that participants with PD would perform worse on the UPDRS, motor tasks, and TMT compared to healthy controls. Second, we hypothesized that measures of number of fixations, fixation duration, pupil dilation, and number of eye blinks would significantly differ between individuals with PD and healthy controls during the IGT. Finally, we hypothesized that overall behavioral performance on the IGT would be worse for our PD group.

## Method and Materials

We secured Institutional Review Board approval through the University of North Texas prior to beginning this study, and participants were recruited through community-based and non-profit organizations, healthcare providers, local businesses, universities, and word of mouth. Prospective participants were given a brief overview of the motor tasks and a general overview of the cognitive assessments used. Examinations occurred in the Neurocognitive laboratory at the University of North Texas.

### Inclusion Criteria for PD Subjects and Matched Healthy Controls

Inclusion criteria included English-speaking/reading/writing individuals between 40-90 years old who self-identified as Black/African American, Non-Hispanic White, and Hispanic with a minimum high school grade level education or General Education Development (GED). A diagnosis of PD was verified during in-person screening with the Movement Disorder Society (MDS) Clinical Diagnostic Criteria from the Unified Parkinson’s Disease Rating Scale (UPDRS). These diagnoses were confirmed by a board-certified neurologist through a private practice as a movement disorder specialist. Groups did not differ between years of education [*t* (43) = 1.82, *p* = .075, *d* = 0.55] or age [*t* (43) = 1.77, p =.083, *d =* 0.53]. Each participant received a $25 gift card as compensation. Exclusion criteria included current autoimmune, metabolic, endocrine, neurological, or psychiatric disorders, self-reported illicit substance use, pregnancy, or substantial visual impairment. Individuals currently taking psychoactive medications were excluded, and participants were asked to abstain from alcohol and tobacco prior to the study. The sample size and primary demographic characteristics of our preliminary participants are listed in Table 1.

**Table 1.** Participant Demographics.

|  | PD Group (N=25) | HC Group (N=20) |
| --- | --- | --- |
| Characteristic | Mean (SD) | Mean (SD) |
| Age (years) | 73.0 (6.3) | 69.6 (6.3) |
| Gender (M/F) | 18/7 | 4/16 |
| Education (years) | 15.2 (2.7) | 16.6 (2.5) |
| Years Since PD Dx | 4.7 (4.4) |  |
| Hoehn and Yahr Stage | 1.44 (.53) |  |

### Demographic/Physiological Assessments

Upon completion of the informed consent, participants completed a basic demographic questionnaire. Blood pressure (systolic/diastolic) and heart rate were then taken, and the Ishihara Test for Color Blindness (48) was administered to screen for color-deficiencies (none were observed).

### UPDRS

The UPDRS was administered to participants to determine stage of PD using the Movement Disorders Society recommended guidelines using four fundamental diagnostic criteria categories: I. Mentation, Behavior, and Mood, II. Activities of Daily Living, III. Motor Examination, and IV. Complications of Therapy (in the past week). Questions were asked relative to each section in parts I-III, while motor function was assessed visually and physically by the investigators in part IV. Parts I-III were scored using a rating scale of 0-4, while part IV uses yes/no indications. Higher scores translate to increased severity (43).

### Motor Tasks

The 6-minute walk test measured travel distance using self-selected gait speed over 6 minutes via stopwatch. Start and stop positions were marked in an empty hallway 30 meters apart using tape. The TUG assesses the amount of time it takes for the participant to get up from a seated position and walk 10 feet at a self-selected gait speed. Start and stop positions were marked using tape.

### Trail Making-Task (TMT-A and B)

Participants performed the TMT A and B test versions using pencil and paper (46). The TMT-A required participants to draw individual lines sequentially connecting 25 encircled numbers randomly positioned on an 8×11 inch test sheet. The TMT-B required participants sequentially connect alternating letters and numbers (e.g., 1, A, 2, B, 3, C, etc.). These tasks were scored based on total completion time and number of errors.

### Barratt’s Impulsiveness Scale (BIS)

Participants were asked 30 Likert-type questions regarding impulsive tendencies with answers ranging from 1 (rarely/never) to 4 (almost always/always) (45). Items were scored based on six primary factors: (attention, motor, self-control, cognitive complexity, perseverance, and cognitive instability) and three second-order factors (attentional, motor, and non-planning). A total composite score was used in analyses with higher scores indicating higher levels of impulsiveness.

### Controlled Oral Word Association test (COWAT)

The COWAT test of verbal fluency included two phases: Animal Naming and FAS (44). For Animal Naming, participants are given 60 seconds to name as many animals as they can think of as quickly as possible. For the FAS test, participants complete three separate 60 second trials where they are asked to orally produce words that begin with the letters F, A, and S as quickly as possible.

### Iowa Gambling Task (IGT)

Participants completed a computerized version of the IGT. Stimuli were presented centrally using E-Prime 3.0 on a 66 cm LCD monitor set to a 1204 x 768-pixel resolution and a 250 Hz refresh rate. The IGT consisted of 100 card selection trials from four identical decks of cards (A, B, C, and D). Participants chose cards one at a time by using a mouse to select their preferred deck. Each time they chose a card, they were given feedback on whether they received a gain or loss of funds based on their choice and “money” was deposited or debited from their account accordingly. All participants started with an amount of $2000 and were told to make a profit. They had no prior knowledge about the amount that the chosen card would yield. Decks A and B always yielded $150, while decks C and D always yielded $50. For each card chosen, there was a 50% chance of having to pay a penalty. For decks A and B, the penalty was $250, whereas for decks C and D it was $50. This task was self-paced and lasted approximately 10 minutes. See Figure 1 for an example.

**Figure 1.**
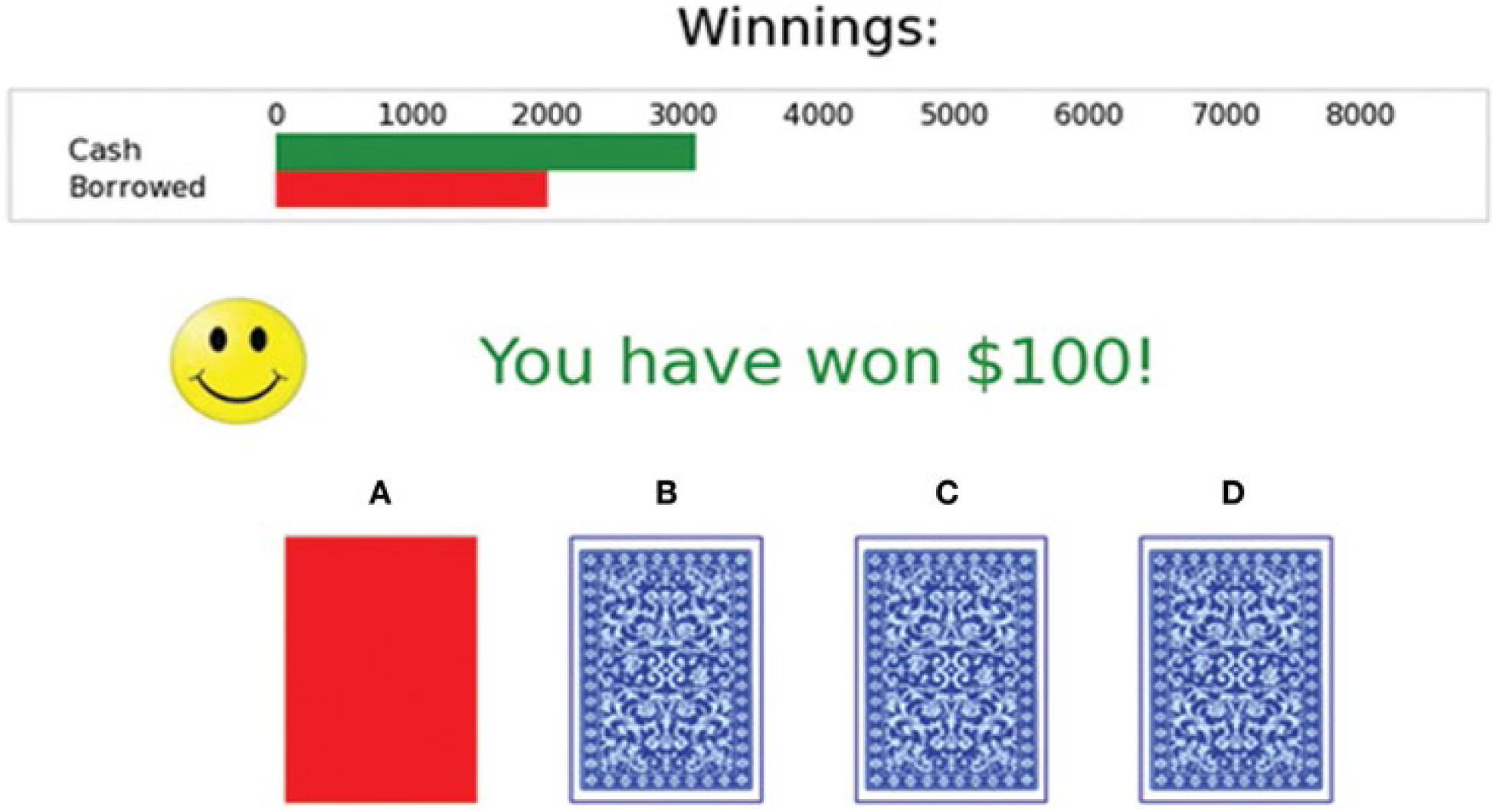
Example of the Computerized IGT with 100 card selection trials from four identical decks of cards (A, B, C, and D). Participants chose cards one at a time by selecting the numbers 1 for deck “A”, 2 for deck “B”, 3 for deck “C”, or 4 for deck “D” on a computer keyboard. Each time they chose a card, they were given feedback on whether they chose correctly or incorrectly, and “money” was deposited (cash) or debited (borrowed) from their account appropriately. All participants started with an amount of $2000 and were told to make a profit. They did not know in advance the amount that the card they chose would yield. Decks A and B always yielded $100, while decks C and D always yielded $50. For each card chosen, there was a 50% chance of having to pay a penalty. For decks A and B, the penalty was $250, whereas for decks C and D it was $50.

### Eye Movements

Eye movements (pupil diameter, fixation, saccades, and blinks) were recorded from the right eye using an Eye Link 1000-Plus tracker with a 250 Hz sampling rate and controlled ambient lighting. Pupil diameter was calculated as mean pupil area in arbitrary camera units (AUs). Although pupil dilation is sometimes reported in metric (mm) units, we chose to preserve our measurement in camera units to avoid any potential transformation artifacts. For reference, 2 mm of pupil dilation corresponds to approximately 435 AU of area and 20.86 AU of diameter with an error rate of less than 1%. Participants were comfortably seated 30 cm (about 11.81 in) from the screen, and their heads were stabilized with a chinrest to minimize movement. Once comfortable, the camera lens was focused, and participants’ eyes were calibrated to the participant’s computer screen (mirrored to the experimenter’s computer screen) and validated using a multipoint grid. After calibration, participants were prompted to begin the IGT, and computerized instructions were given while a researcher remained in the testing room. Eye movements were recorded continuously throughout the task. For analysis, averages for each type of eye movement were taken for the full trial period (100 trials total).

## Results

### Statistical Analyses

Performance on UPDRS, motor assessments, TMT A and B, BIS, COWAT, IGT, and eye movement measures were analyzed using a series of one-way ANOVAs to test for between-groups differences. We also conducted a general linear model on block-by-block performance over time in the IGT to assess learning and decision-making over time. For each outcome variable of interest, we conducted maximum normalized residual analyses to identify extreme values and report as outliers that could bias results (49). We also used exploratory partial least squares correlations between our eye fixation duration results and IGT blocks to shed light on learning behaviors. For all analyses, alpha was set at p < .05, and effect sizes are reported.

### Unified Parkinson’s Disease Rating Scale (UPDRS)

To assess the differences in Parkinson’s disease symptoms among the two groups (PD group and healthy control group), we used summarized subscale scores from the three sections of the Unified Parkinson’s Disease Rating Scale (UPDRS): Part I (Mentation, Behavior, and Mood), Part II (Activities of Daily Living), and Part III (Motor Examination). As expected, results revealed significant group effects for each of the three subscales with PD patients scoring higher for each outcome measure (listed in Table 2 below).

**Table 2.**
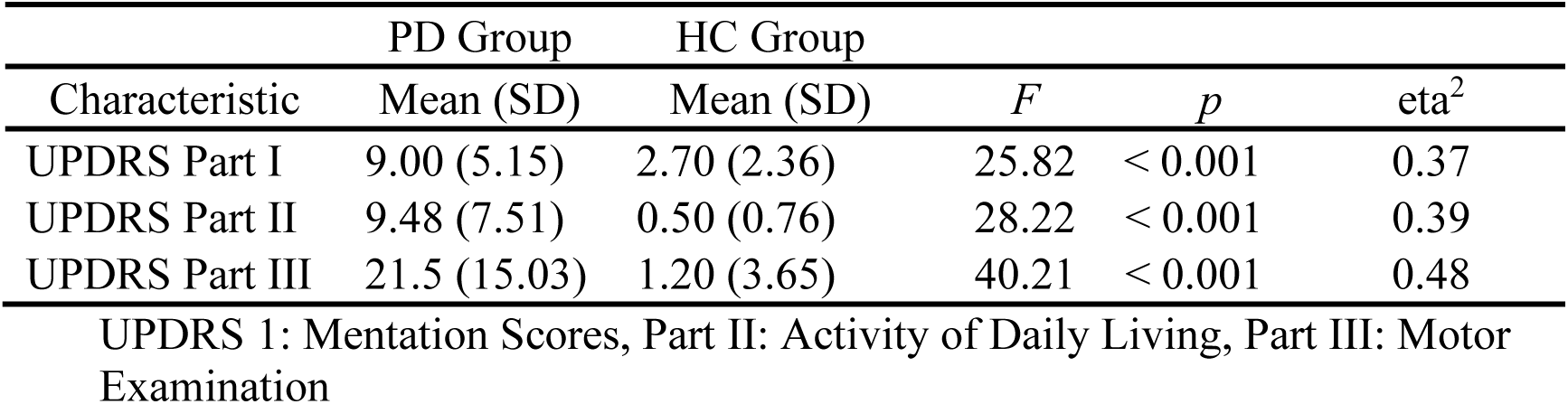
UPDRS Between-Groups Comparisons.

### Motor Assessments

Distance and speed were analyzed for the three different motor function assessments (6-minute Walk Test, TUG test, and Gait Speed) (Listed in Table 3 below). Results revealed a significant group difference indicating that individuals with PD walked a shorter duration than healthy controls. For the TUG analysis, four individuals were identified as outliers due to scoring difficulties with timing errors. Therefore, as these outliers were excluded from the analyses, this comparison was conducted on *N*=41 (21 PD, 20 Matched Controls). Comparisons for TUG and Gait Speed analyses revealed no significant between-groups differences (see Table 3).

**Table 3.**
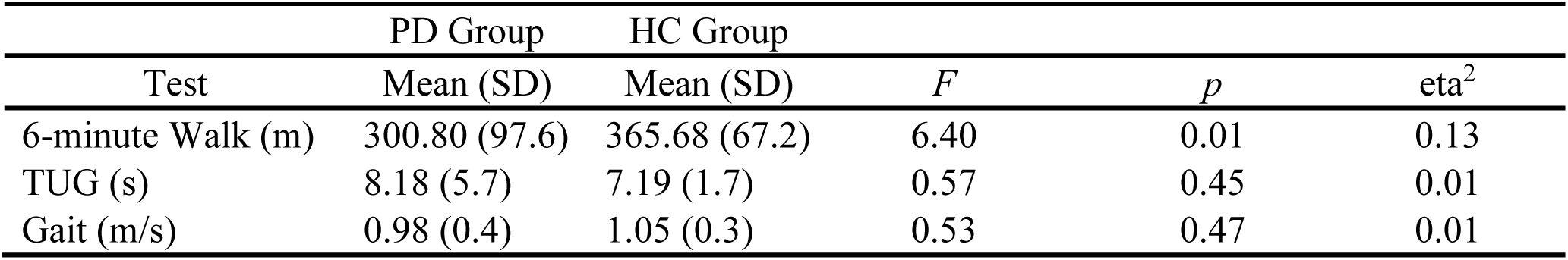
Motor Results.

### Cognitive Assessments

#### Trail Making Test (TMT A & B)

Results for all cognitive assessments are listed in Table 4. Outlier analyses identified two extreme values for TMT A and TMT B in the PD group, thus results are reported for *N*=23 and *N*=20, respectively. Between-groups differences in mean completion times (seconds) were significant for both TMT A and TMT B, with healthy controls completing both tests faster than the PD group. The average number of errors did not differ between groups for version A. However, PD participants exhibited a significantly higher number of errors than controls in version B. This indicates that those with early-stage PD need a longer amount of time to process information that requires shifting attention –even with familiar letters and numbers.

**Table 4.**
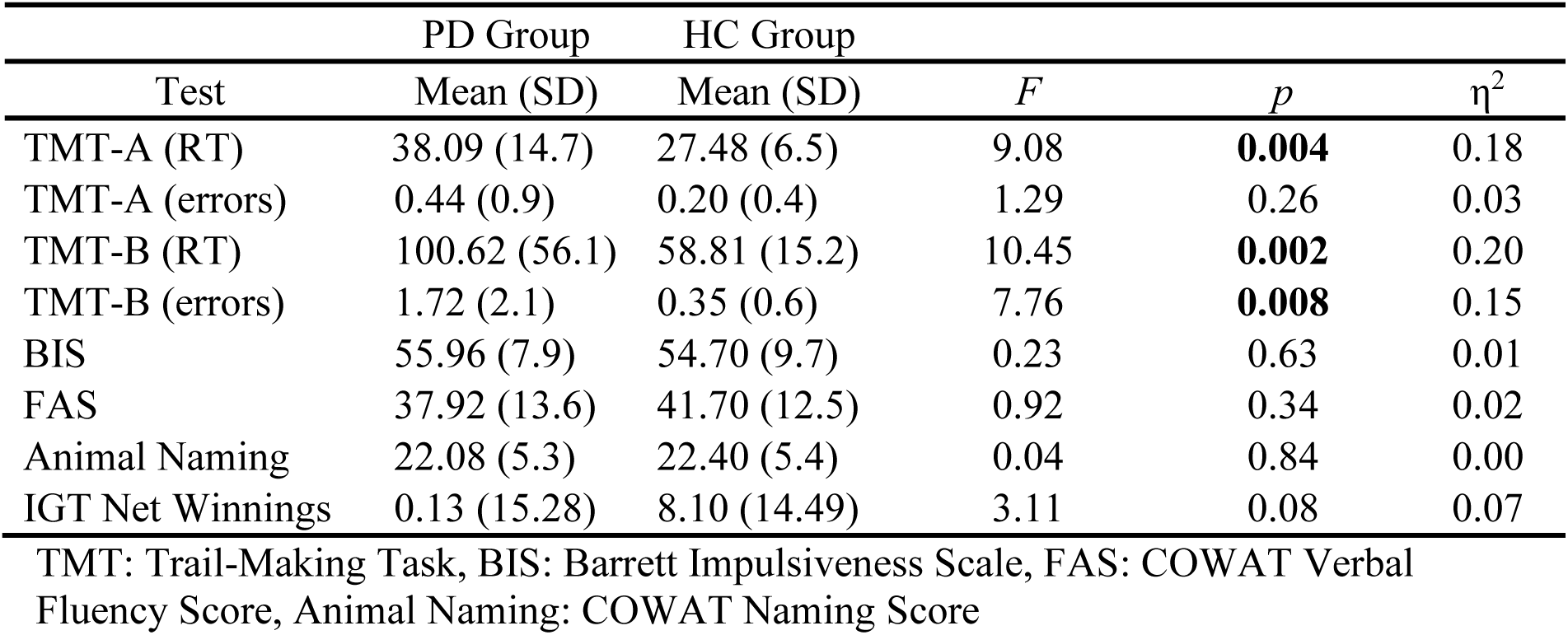
Cognitive and IGT Test Results.

#### Barratt Impulsiveness Scale (BIS)

For the BIS, a between-groups comparison was not statistically significant, indicating that the PD group performed similarly to healthy controls.

#### Controlled Oral Word Association Test (COWAT)

The total number of words produced for letters F, A, and S were summed to create a composite verbal fluency score, while the Animal Naming was used to assess semantic memory. No significant between groups differences were observed for either verbal fluency or semantic fluency.

#### Iowa Gambling Task (IGT)

Outlier analyses identified one extreme value for net winning scores on the IGT in the PD group, thus analyses were conducted on N=24 PD and N=20 Controls. Results revealed a group effect approached significance [*F* (43) *=* 3.11, *p* =.08], suggesting that the PD group accumulated more losses than controls. To investigate IGT performance on a more precise level, we conducted a general linear model (GLM) across the task broken up into five 20-trial blocks to determine deck-specific responding over time that could reveal problematic learning behaviors. Results indicated a significant main effect of block [*F* (3, 39) *=* 3.08, *p* =.03, _p_η^2^ = .24]. Interestingly, this was qualified by a block by group interaction, [*F* (4, 39) *=* 2.77, *p* =.04, _p_η^2^ = .22]. Specifically, according to net winnings by the end of the task (block 5), PD participants were persistently making disadvantageous as opposed to advantageous choices compared to healthy controls [*F* (43) *=* 7.69, *p* =.008 _p_η^2^ = .15] (see figures 2 and 3).

**Figure 2.**
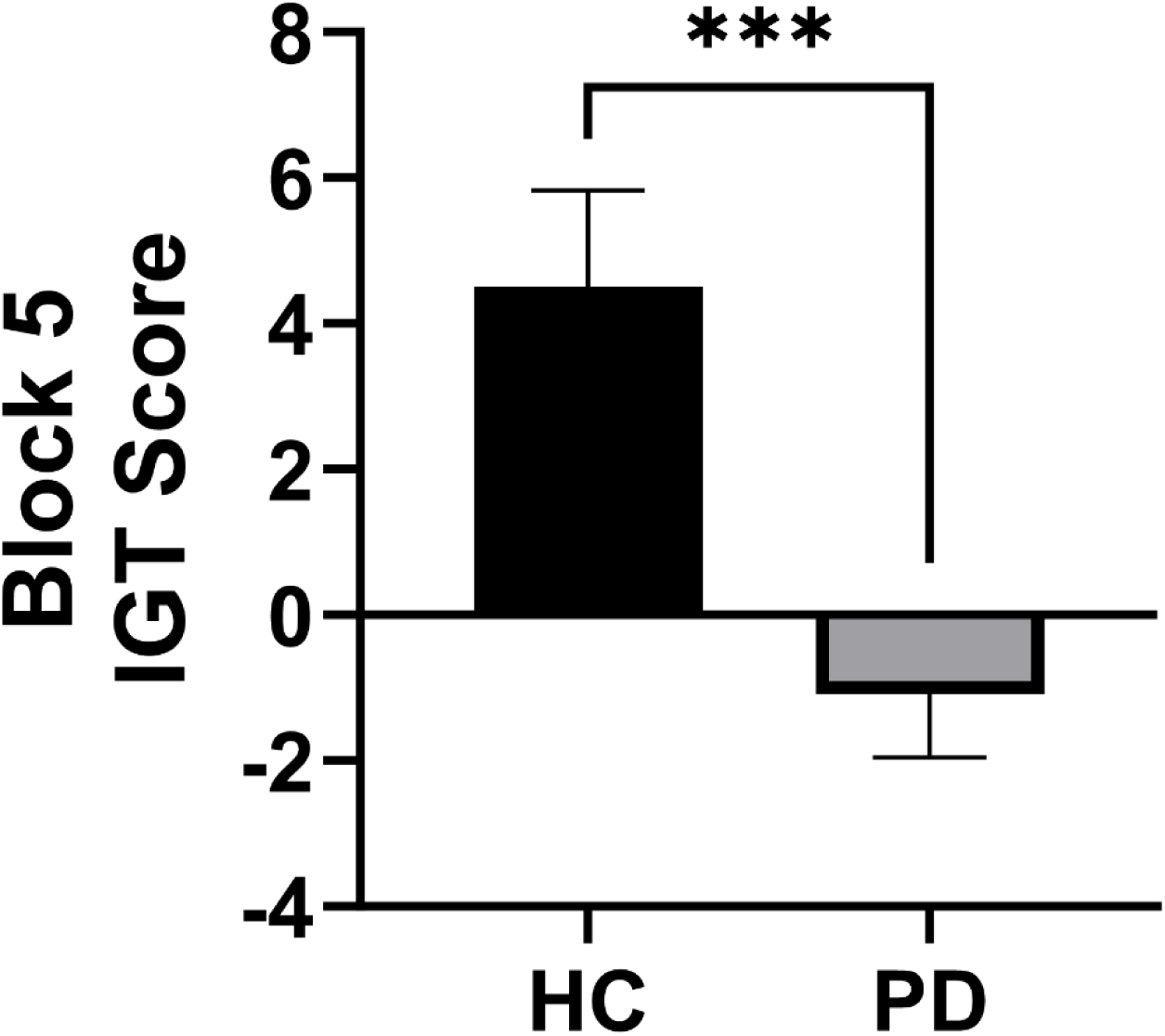
Block 5 net winning scores for the Ioway Gambling Task (IGT) comparing participants with Parkinson’s (PD) to Healthy Controls (HC) by the end of the task (*p*=.008).

**Figure 3.**
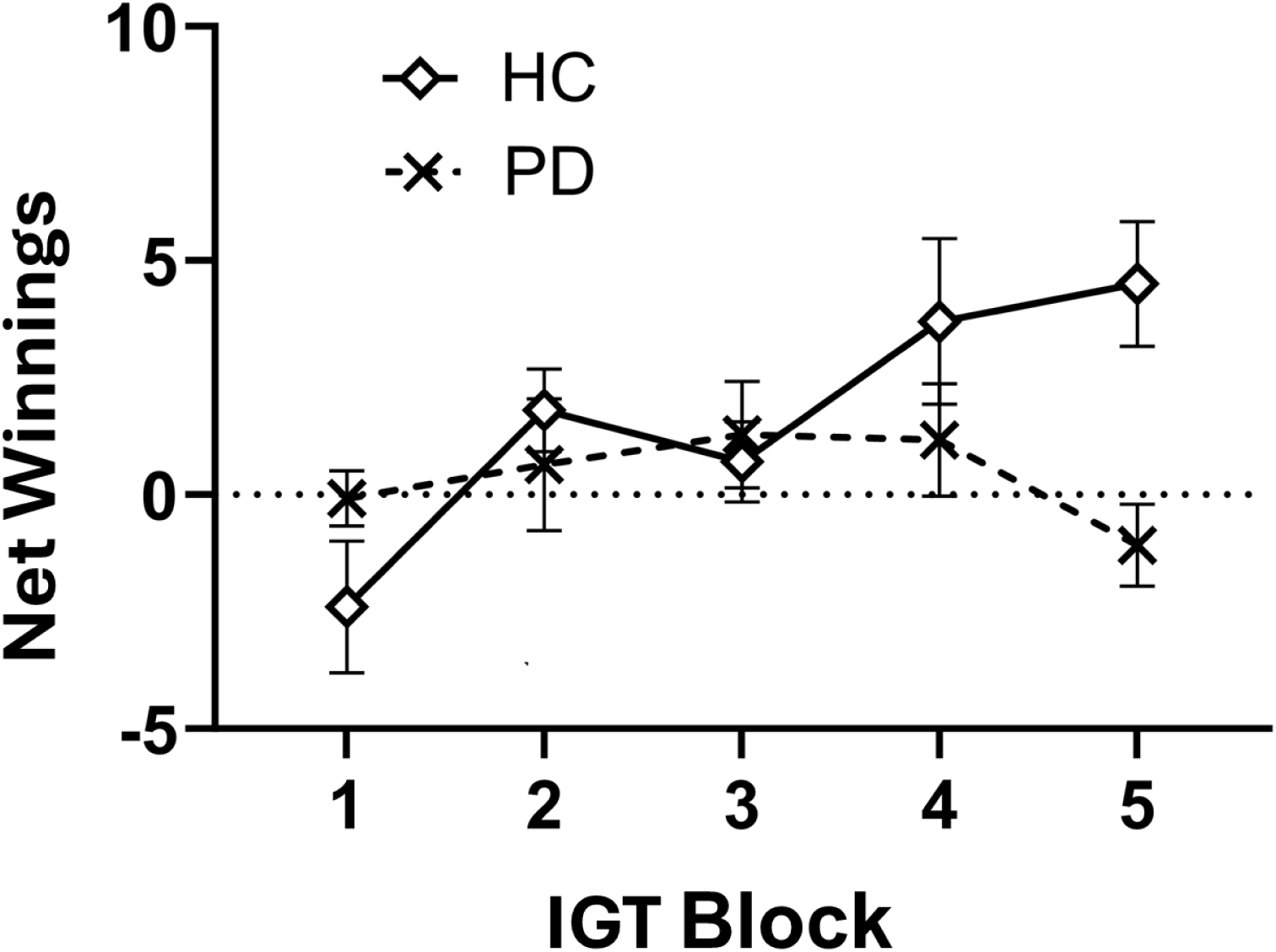
Net winning scores for the Iowa Gambling Task (IGT) analyzed as a function of performance over time in five 20-trial blocks (100 trials total). By block 5, results suggest individuals with Parkinson’s (PD) maintained a disadvantageous selection strategy by the end of the task compared to Healthy Controls (HC).

#### Eye Movements During the IGT

Eye movement results are listed in Table 5. Recall that we analyzed several eye tracking metrics: average fixation count, average fixation duration, average saccade count, average blink count, and average pupil dilation during the entirety of the IGT. The mean trial completion times for individual trials were highly variable in the present study given the self-paced nature of the task (Range = 904.80-8818.30 ms). We therefore analyzed our eye movement data based upon mean participant responses across trials.

**Table 5.** Eye Movement Results.

| Eye Movement | PD Group | HC Group | F | *p* | $\eta^2$ |
| --- | --- | --- | --- | --- | --- |
|  | Mean (SD) | Mean (SD) |  |  |  |
| Fixation Count | 5.80 (2.5) | 5.17 (2.3) | 0.78 | 0.381 | 0.02 |
| Fixation Duration | 336.37 (92.0) | 308.94 (114.8) | 0.79 | 0.390 | 0.02 |
| Saccade Count | 5.05 (2.6) | 4.46 (2.1) | 0.69 | 0.409 | 0.02 |
| Blink Count | 0.19 (0.3) | 0.67 (0.4) | 22.37 | **<0.001** | 0.37 |
| Pupil Dilation | 2772.14<br>(376.8) | 2869.36<br>(367.7) | 0.76 | 0.378 | 0.02 |

Eye movements represent the mean number of observations averaged across trials for the IGT Fixation duration is reported in milliseconds, Pupil dilation is reported in arbitrary units (au)

#### Fixation Count

Groups did not differ significantly, thus suggesting that PD participants and controls exhibited an equivalent number of fixations during the IGT.

#### Fixation Duration

Similarly, average fixation duration did not differ significantly between groups during the IGT. Based upon our observation of a numerical trend, we conducted a series of exploratory partial correlations for fixation durations during each of the five IGT blocks controlling for group. While fixation durations were not significantly correlated with performance in blocks 1 or 4 [rPLS (42) = −0.16, p=0.30; rPLS (42) = 0.03, p=0.81, respectively], they *<u>were</u>*significantly correlated with block 2 [rPLS (42) = 0.35, p =0.02], block 3 [rPLS (42) = 0.34, p =0.02], and block 5 [rPLS (42) = 0.37, p=0.01]. These extraordinary novel findings suggest a unique and revealing relationship between eye fixation durations and the ability (or inability) to learn and implement advantageous decision-making strategies.

#### Saccade Count

Group differences in saccades were not significant, thus suggesting that PD participants and controls exhibited an equivalent number of saccadic movements during the IGT.

#### Blink Count

An analysis of average blink count revealed a highly significant difference between groups. Individuals with PD exhibited a 72% reduction in eye blinks during the IGT compared to the healthy control group, resulting in a very large effect size. Like our results with fixation duration, the blink count is an extremely valuable outcome that underscores the role of striatal dopamine function in PD, given that this anatomical area and neurochemical are involved in reinforcement-based learning over time.

#### Pupil Dilation

Group differences in mean pupil dilation (measured in area) did not differ significantly, suggesting an equivalent level of dilation-related working memory between PD participants and controls.

## Discussion

In support of our study goals, we found significant relationships and group differences using the IGT and eye movement abnormalities in PD subjects vs. controls. Using a novel combination of the IGT with eye tracking and additional cognitive and motor tests, we compared early-stage to mid-stage individuals with PD to matched healthy controls. In support of our first hypothesis, participants with PD scored significantly higher on each of the three UPDRS subscales than controls, and completion times on the TMT A and B were significantly longer in the PD group compared to controls. Additionally, participants in the PD group exhibited a higher number of errors on the TMT B compared to their control counterparts. This pattern is consistent with established norms (50), and group differences are consistent with previous findings of impaired executive function in PD (13–15, 51). Motor performance during the TUG and Gait Speed tests did not differ between groups, however, participants in the PD group traveled a significantly shorter distance than the healthy control group during the six-minute walk test. Our measures of impulsivity (BIS-11), and verbal fluency (COWAT) revealed similar scores between groups. There was no significant difference in TUG test results between groups. While many findings report differences in TUG scores in PD, a pattern of equivalent scores between early PD and controls on this measure has precedence (52). Similarly, while previous findings often reflect group differences in BIS and COWAT scores, our findings could suggest that our PD sample had not yet reached diagnostic thresholds for impulse control disorders (53) or impoverished oral semantic fluency (54).

Consistent with our second hypothesis, we did observe a highly significant group effect for eye blinks during the IGT. Surprisingly, PD participants exhibited a 70% reduction in blinks compared to controls. This is in accordance with two published studies from Japan reporting a decrease in task-independent spontaneous blink rates in mid-stage PD patients (55,56). Contrary to our predictions, group differences for average number of saccades, fixations, or fixation duration values or pupil dilation values a did not differ between group as expected. Exploratory analyses did reveal significant correlations between fixation duration and performance in the middle and end blocks of the IGT, although these results should be interpreted with caution.

The difference in overall net IGT winning scores approached significance between groups. Upon further consideration of potential changes in decision-making over time in accordance with previous work (e.g., 24), we discovered that by the fifth and final block of trials, individuals with PD had achieved negative net winning scores (i.e., losses) whereas the healthy controls had accumulated positive net winnings. This is consistent with our third hypothesis. The current literature on IGT in Parkinson’s is mixed with respect to behavioral performance (20). Some previous studies have reported reduced performance in PD compared to healthy controls, and a handful have reported no differences (e.g., 57). The present results support the utility of the IGT for assessing behavioral changes in early PD and underscore the importance of subtleties in IGT performance early in may not be fully captured by overall performance, as evidenced by changes over time in the task. When considering decision-making over time during the IGT, our findings also indicate that PD subjects maintained a disadvantageous strategy for deck selection, perhaps reflecting an insensitivity to aversive outcomes in reward-based learning (20,23,24). This is an accordance with the hypothesis that dopaminergic changes in PD involving substantia nigra, basal ganglia, and prefrontal cortex functionally manifest as an insensitivity to feedback (in this case an inability to capitalize on negative feedback critical for error correction). The IGT also assesses cognitive flexibility by simulating real-world decision-making under conditions of uncertainty (17). In a recent comprehensive review, Salvatore and colleagues propose that the IGT may be valuable for evaluating possible PD pathology in the prodromal phase or propensity for therapeutic adherence (20). Specifically, non-strategic decision-making may correlate with non-compliance, or self-discontinuing, therapeutic regimens. Although there is an increased risk for life-threatening complications as the disease progresses, up to 70% of PD patients still choose to self-discontinue therapy (7,8). This importantly reveals a PD patient’s inability to assess and weigh consequences in critical decision making.

With respect to our eye movement measures, we demonstrate that older adults with PD exhibit substantially fewer spontaneous eye blinks on average compared to healthy older adults during the IGT. This likely reflects impaired dopaminergic and norepinephrinergic function involved in both cognition and motor function (31–34). Additionally, decreased dopaminergic activity may lead to Impaired motor execution in targeted regions innervated by the corticobulbar tract. This pathway is involved in regulating movements of the face, head, and neck by influencing the cranial nerves, including those that control the extraocular muscles (cranial nerves III, IV, and VI). Although it does not directly coordinate eye blinks, it contributes indirectly to voluntary eye movement through its interaction with brainstem regions. A reduction in blink rate may contribute to the hallmark hypomimia or “facial masking” often present in PD, a phenomenon with psychosocial implications for individuals with PD and caretakers (58). This finding in particular offers strong support for the utility of eyeblinks as a diagnostic marker during decision-making and other cognitive tasks (31,59). An important research point to consider is that eyeblinks are often considered artifacts and are thus removed from eye tracking recordings during data cleaning and extraction. Researchers should reconsider the value of these data, especially given the link to dopaminergic function.

This is the first study, to our knowledge, to combine eye tracking with the IGT in individuals with PD. These novel results suggest that while behavioral performance on the IGT may not always differ on a gross scale between healthy older adults and those with early to mid-stage PD, there are nuanced and subtle distinctions, particularly with respect to shifts in strategies over time, that distinguish the disease. Combined with non-invasive psychophysiological measures such as eye tracking, the IGT and similar cognitive tasks may be of clinical utility (10).

Our study is not without limitations. Participants represented a well-educated, high-functioning, and racially/ethnically homogeneous subset of the population with access to medical care. Future research should consider balancing racial/ethnic and gender demographics. Also, given the rather small sample size in the present study, it is possible that our experimental power was limited to detect differences in fixations, saccades, and pupil dilation during eye movements. Our brief measure of impulsivity may not have been sensitive enough, thus future work should aim to incorporate Parkinson’s specific measures of impulse control disorders (53). Finally, adapting a standardized version of the IGT for eye tracking would be beneficial for future research allowing for fine-grained analyses of discrete windows of oculomotor movements during different phases of the task (e.g., initial presentation, deck selection, feedback). The relatively short duration of each trial and the variability of self-paced response timing in the present study was limiting for this type of analysis, and this would be an excellent direction through which researchers could extend these findings.

In conclusion, IGT holds promise for assessing cognitive changes in PD particularly for evaluating decision-making abilities. As PD progresses, subtle changes in cognition, such as executive function, decision-making, and risk assessment, may decline. This has key implications for daily life and treatment decisions. Considering the currently projected doubling of individuals who will develop PD in their lifetime (60), combining sensitive measures such as eye tracking, with cognitive testing and other disease markers may be an important and simple way to cognitive detect dysfunction early in PD to better characterize disease progression and hasten intervention.

## Data Availability

All data produced in the present study are available upon reasonable request to the authors

## Author Contributions

KD and AR wrote the manuscript; KD conducted the experiments; KD, AR, VN, and MS assisted with conceptual organization, data analyses, and editing of the manuscript. We thank Dr. Jerome Lisk for his assistance in recruitment and guidance with PD diagnostics. Finally, we would like to thank Antonio Archuleta, Natalee Sidorchuk, and Prisha Goyal for assistance with data collection and coding.

## References

1. Kordower JH, Olanow CW, Dodiya HB, Chu Y, Beach TG, Adler CH, Halliday GM, Bartus RT. Disease duration and the integrity of the nigrostriatal system in Parkinson’s disease. Brain. 2013 Aug 1;136(8):2419–31.

2. Kouli A, Torsney KM, Kuan WL. Parkinson’s disease: etiology, neuropathology, and pathogenesis. Exon Publications. 2018 Dec 21:3–26.

3. Wirdefeldt K, Adami HO, Cole P, Trichopoulos D, Mandel J. Epidemiology and etiology of Parkinson’s disease: a review of the evidence. European journal of epidemiology. 2011 Jun; 26:1–58.

4. McDowell K, Chesselet MF. Animal models of the non-motor features of Parkinson’s disease. Neurobiology of disease. 2012 Jun 1;46(3):597–606.

5. Durcan R, Wiblin L, Lawson RA, Khoo TK, Yarnall AJ, Duncan GW, Brooks DJ, Pavese N, Burn DJ, ICICLE-PD Study Group. Prevalence and duration of non-motor symptoms in prodromal Parkinson’s disease. European journal of neurology. 2019 Jul;26(7):979–85.

6. Goldman JG, Litvan I. Mild cognitive impairment in Parkinson’s disease. Minerva medica. 2011 Dec;102(6):441.

7. Straka I, Minar M, Skorvanek M. Adherence to pharmacotherapy in patients with Parkinson’s disease taking three and more daily doses of medication. Front Neurol. 2019; 10: 799.

8. Daley DJ, Myint PK, Gray RJ, Deane KH. Systematic review on factors associated with medication non-adherence in Parkinson’s disease. Parkinsonism & related disorders. 2012 Dec 1;18(10):1053–61.

9. Fengler S, Liepelt-Scarfone I, Brockmann K, Schäffer E, Berg D, Kalbe E. Cognitive changes in prodromal Parkinson’s disease: a review. Movement Disorders. 2017 Dec;32(12):1655–66.

10. Antoniades, C. A., & Spering, M. Eye movements in Parkinson’s disease: from neurophysiological mechanisms to diagnostic tools. Trends in Neurosciences. 2014 Jan; 47(1): 71–83.

11. Nejtek VA, James RN, Salvatore MF, Alphonso HM, Boehm GW. Premature cognitive decline in specific domains found in young veterans with mTBI coincide with elder normative scores and advanced-age subjects with early-stage Parkinson’s disease. 2021 PLoS ONE 16(11): e0258851.

12. Friedman NP, Miyake A. Unity and diversity of executive functions: Individual differences as a window on cognitive structure. Cortex. 2017 Jan 1; 86:186–204.

13. Aron AR, Robbins TW, Poldrack RA. Inhibition and the right inferior frontal cortex. Trends in cognitive sciences. 2004 Apr 1;8(4):170–7.

14. Dulas MR, Duarte A. Aging affects the interaction between attentional control and source memory: an fMRI study. Journal of cognitive neuroscience. 2014 Dec 1;26(12):2653–69.

15. Dulas MR, Duarte A. Age-related changes in overcoming proactive interference in associative memory: The role of PFC-mediated executive control processes at retrieval. Neuroimage. 2016 May 15; 132:116–28.

16. Botvinick MM, Cohen JD, Carter CS. Conflict monitoring and anterior cingulate cortex: an update. Trends in cognitive sciences. 2004 Dec 1;8(12):539–46.

17. Bechara A, Damasio AR. The somatic marker hypothesis: A neural theory of economic decision. Games and economic behavior. 2005 Aug 1;52(2):336–72.

18. Perugini A, Ditterich J, Shaikh AG, Knowlton BJ, Basso MA. Paradoxical decision-making: a framework for understanding cognition in Parkinson’s disease. Trends in neurosciences. 2018 Aug 1;41(8):512–25.

19. Colautti L, Iannello P, Silveri MC, Antonietti A. Decision making in Parkinson’s disease: an analysis of the studies using the Iowa gambling task. European Journal of Neuroscience. 2021 Nov;54(10):7513–49.

20. Salvatore MF, Soto I, Alphonso H, Cunningham R, James R, Nejtek VA. Is there a neurobiological rationale for the utility of the Iowa Gambling Task in Parkinson’s disease? Journal of Parkinson’s Disease. 2021 Jan 1;11(2):405–19.

21. Castrioto A, Funkiewiez A, Debû B, Cools R, Lhommée E, Ardouin C, Fraix V, Chabardès S, Robbins TW, Pollak P, Krack P. Iowa gambling task impairment in Parkinson’s disease can be normalised by reduction of dopaminergic medication after subthalamic stimulation. Journal of Neurology, Neurosurgery & Psychiatry. 2015 Feb 1;86(2):186–90.

22. Xi C, Zhu Y, Mu Y, Chen B, Dong B, Cheng H, Hu P, Zhu C, Wang K. Theory of mind and decision-making processes are impaired in Parkinson’s disease. Behavioural Brain Research. 2015 Feb 15; 279:226–33.

23. Kobayakawa M, Tsuruya N, Kawamura M. Sensitivity to reward and punishment in Parkinson’s disease: an analysis of behavioral patterns using a modified version of the Iowa gambling task. Parkinsonism & Related Disorders. 2010 Aug 1;16(7):453–7.

24. Kobayakawa M, Tsuruya N, Kawamura M. Decision-making performance in Parkinson’s disease correlates with lateral orbitofrontal volume. Journal of the neurological sciences. 2017 Jan 15; 372:232–8.

25. Alberico SL, Cassell MD, Narayanan NS. The vulnerable ventral tegmental area in Parkinson’s disease. Basal Ganglia. 2015 Aug 1;5(2-3):51–55.

26. Mather M, Harley CW. The locus coeruleus: essential for maintaining cognitive function and the aging brain. Trends in cognitive sciences. 2016 Mar 1;20(3):214–26.

27. Spering M. Eye movements as a window into decision-making. Annual review of vision science. 2022 Sep 15;8(1):427–48.

28. McGinty VB, Rangel A, Newsome WT. Orbitofrontal cortex value signals depend on fixation location during free viewing. Neuron. 2016 Jun 15;90(6):1299–311.

29. Ding J, Powell D, Jiang Y. Dissociable frontal controls during visible and memory-guided eye-tracking of moving targets. Human brain mapping. 2009 Nov;30(11):3541–52.

30. Hui M, Beier KT. Defining the interconnectivity of the medial prefrontal cortex and ventral midbrain. Front Mol Neurosci. 2022 Jul 22;(15):971349.

31. Van Slooten JC, Jahfari S, Theeuwes J. Spontaneous eye blink rate predicts individual differences in exploration and exploitation during reinforcement learning. Scientific reports. 2019 Nov 22;9(1):17436

32. Jongkees BJ, Colzato LS. Spontaneous eye blink rate as predictor of dopamine-related cognitive function—A review. Neuroscience & Biobehavioral Reviews. 2016 Dec 1; 71:58–82.

33. Willett SM, Maenner SK and Mayo JP. The perceptual consequences and neurophysiology of eye blinks. Front. Syst. Neurosci. 2023 Aug 17:1242654

34. Redgrave, P., Rodriguez, M., Smith, Y. et al. Goal-directed and habitual control in the basal ganglia: implications for Parkinson’s disease. Nat Rev Neurosci.2010 Oct 11:760–772.

35. Eckstein MK, Guerra-Carrillo B, Singley AT, Bunge SA. Beyond eye gaze: What else can eyetracking reveal about cognition and cognitive development? Developmental cognitive neuroscience. 2017 Jun 1; 25:69–91.

36. Zhou C, Guo T, Bai X, Wu J, Gao T, Guan X, Liu X, Gu L, Huang P, Xuan M, Gu Q. Locus coeruleus degeneration is associated with disorganized functional topology in Parkinson’s disease. NeuroImage: Clinical. 2021 Jan 1; 32:102873.

37. Wong OW, Chan AY, Wong A, Lau CK, Yeung JH, Mok VC, Lam LC, Chan S. Eye movement parameters and cognitive functions in Parkinson’s disease patients without dementia. Parkinsonism & related disorders. 2018 Jul 1; 52:43–8.

38. Archibald NK, Hutton SB, Clarke MP, Mosimann UP, Burn DJ. Visual exploration in Parkinson’s disease and Parkinson’s disease dementia. Brain. 2013 Mar 1;136(3):739–5.

39. Wang CA, McInnis H, Brien DC, Pari G, Munoz DP. Disruption of pupil size modulation correlates with voluntary motor preparation deficits in Parkinson’s disease. Neuropsychologia. 2016 Jan 8; 80:176–84.

40. Tsitsi P, Benfatto MN, Seimyr GÖ, Larsson O, Svenningsson P, Markaki I. Fixation duration and pupil size as diagnostic tools in Parkinson’s disease. Journal of Parkinson’s Disease. 2021 Jan 1;11(2):865–75.

41. Ranchet M, Orlosky J, Morgan J, Qadir S, Akinwuntan AE, Devos H. Pupillary response to cognitive workload during saccadic tasks in Parkinson’s disease. Behavioural brain research. 2017 Jun 1; 327:162–6.

42. Hoehn MM, Yahr MD. Parkinsonism: onset, progression, and mortality. Neurology. 1967 May;17(5):427.

43. Goetz CG, Tilley BC, Shaftman SR, Stebbins GT, Fahn S, Martinez-Martin P, Poewe W, Sampaio C, Stern MB, Dodel R, Dubois B. Movement Disorder Society-sponsored revision of the Unified Parkinson’s Disease Rating Scale (MDS-UPDRS): scale presentation and clinimetric testing results. Movement disorders: official journal of the Movement Disorder Society. 2008 Nov 15;23(15):2129–70.

44. Benton AL, Hamsher DS, Sivan AB. Controlled oral word association test. Archives of Clinical Neuropsychology. 1994.

45. Patton JH, Stanford MS, Barratt ES. Factor structure of the Barratt impulsiveness scale. Journal of clinical psychology. 1995 Nov;51(6):768–74.

46. Reitan RM, Tarshes EL. Differential effects of lateralized brain lesions on the Trail Making Test. The Journal of nervous and mental disease. 1959 Sep 1;129(3):257–62.

47. Bechara A, Damasio AR, Damasio H, Anderson SW. Insensitivity to future consequences following damage to human prefrontal cortex. Cognition. 1994 Apr;50(1-3):7–15.

48. Clark JH. The Ishihara test for color blindness. American Journal of Physiological Optics. 1924.

49. Grubbs FE. Procedures for detecting outlying observations in samples. Technometrics. 1969 Feb 1;11(1):1–21.

50. Tombaugh TN. Trail Making Test A and B: normative data stratified by age and education. Archives of clinical neuropsychology. 2004 Mar 1;19(2):203–14.

51. Kudlicka A, Clare L, Hindle JV. Executive functions in Parkinson’s disease: Systematic review and meta-analysis. Movement disorders. 2011 Nov;26(13):2305–15.

52. Zampieri C, Salarian A, Carlson-Kuhta P, Aminian K, Nutt JG, Horak FB. The instrumented timed up and go test: potential outcome measure for disease modifying therapies in Parkinson’s disease. Journal of Neurology, Neurosurgery & Psychiatry. 2010 Feb 1;81(2):171–6.

53. Giovannelli F, Gavazzi G, Noferini C, Palumbo P, Viggiano MP, Cincotta M. Impulsivity Traits in Parkinson’s Disease: A Systematic Review and Meta-Analysis. Movement Disorders Clinical Practice. 2023 Oct;10(10):1448–58.

54. Hedman E, Hartelius L, Saldert C. Word-finding difficulties in Parkinson’s disease: Complex verbal fluency, executive functions and other influencing factors. Int J Lang Commun Disord. 2022 May;57(3):565–577.

55. Iwaki H, Sogo H, Morita H, Nishikawa N, Ando R, Miyaue N, Tada S, Yabe H, Nagai M, Nomoto M. Using Spontaneous Eye-blink Rates to Predict the Motor Status of Patients with Parkinson’s Disease. Intern Med. 2019;58(10):1417–1421.

56. Kimura N, Watanabe A, Suzuki K, Toyoda H, Hakamata N, Fukuoka H, Washimi Y, Arahata Y, Takeda A, Kondo M, Mizuno T, Kinoshita S. Measurement of spontaneous blinks in patients with Parkinson’s disease using a new high-speed blink analysis system. J Neurol Sci. 2017 Sep 15; 380:200–204.

57. Poletti M, Frosini D, Lucetti C, Del Dotto P, Ceravolo R, Bonuccelli U. Iowa Gambling Task in de novo Parkinson’s disease: a comparison between good and poor performers. Movement Disorders. 2012 Feb;27(2):330–2.

58. Bianchini E, Rinaldi D, Alborghetti M, Simonelli M, D’Audino F, Onelli C, Pegolo E, Pontieri FE. The Story behind the Mask: A Narrative Review on Hypomimia in Parkinson’s Disease. Brain Sciences. 2024 Jan 22;14(1):109.

59. Tao L, Wang Q, Liu D, Wang J, Zhu Z, Feng L. Eye tracking metrics to screen and assess cognitive impairment in patients with neurological disorders. Neurological Sciences. 2020 Jul;41:1697–704.

60. Obeso JA, Stamelou M, Goetz CG, Poewe W, Lang AE, Weintraub D, Burn D, Halliday GM, Bezard E, Przedborski SJ, Lehericy S. Past, present, and future of Parkinson’s disease: A special essay on the 200th Anniversary of the Shaking Palsy. Movement disorders. 2017 Sep;32(9):1264–310.

